# Fully automated segmentation of the locus coeruleus: application to assess the effects of age, sex and education

**DOI:** 10.1101/2025.01.08.25320149

**Authors:** Robin de Flores, Tao Blanchard, Emilie Foyard, Mikaël Naveau, Nicolas Delcroix, Léa Chauveau, Brigitte Landeau, Denis Vivien, Paul A. Yushkevich, Gaël Chételat, Medit-Ageing research group

## Abstract

**Background:** Locus coeruleus (LC) imaging using neuromelanin-sensitive (NM) MRI sequences is a promising biomarker for detecting early Alzheimer’s Disease (AD) and Parkinson’s Disease (PD). Although semi or fully automatic approaches have been developed to estimate LC integrity by measuring its intensity, these techniques most often rely on a single template built in a standardized space and/or depend on a number of voxels to be accounted that is defined a priori. Thus, these algorithms make it impossible to perform direct volumetric analyses and do not properly account for inter-individual anatomical variability. To fill this gap, our aim was to develop a new multi-atlas fully automated segmentation method using the *Automatic Segmentation of Hippocampal Subfields* (ASHS) software. A second aim was to investigate the effects of age, sex and education on LC intensity and volume.

**Method:** We used cross-sectional data from 101 cognitively unimpaired older adults (mean age: 73.8±6.6 years; mean education: 13.2±3.0 years; 58 women, 43 men) from the Age-Well randomized controlled trial for whom high-resolution NM-MRI (T1-w with magnetization transfer; 0.3×0.3×0.75mm^3^) and standard T1-w MRI (1×1x1mm^3^) were available. The LC were manually segmented in 30 randomly selected participants on NM-MRI, and the standard T1-w MRI, NM-MRI and bilateral segmentations were fed into the ASHS training pipeline to generate a new atlas (ASHS-LC). ASHS-LC was applied to the 71 remaining subjects to segment the LC and we assessed the effects of age, sex and education on both i) LC intensity (normalized by the intensity of the pons) and ii) LC volume (normalized by the total intracranial volume).

**Result:** Five-fold cross-validation experiments revealed high accuracy of the automatic segmentation relative to manual segmentation (Dice coefficient 0,83±0,04). ICCs demonstrated excellent reliability for intensity (ICC = 0.99), whereas reliability for volume was lower (ICC = 0.43). LC intensity was significantly higher in women than in men (Cohen’s d = 0.94, p < 0.001) while no associations with age (β = -0.0002, p = 0.98) or education (β = 0.11, p = 0.37) were found. In contrast, LC volume was not different between men and women (Cohen’s d = 0.10, p= 0.55) but was significantly negatively associated with age (β = -0.25, p = 0.04) and education (β = -0.27, p = 0.04). Lastly, LC intensity and volume were not statistically correlated, neither using manual segmentation (atlas set: β = 0.05, p = 0.78) nor ASHS segmentation (atlas set: β = 0.11, p = 0.57; analysis set: β = -0.07, p = 0.55).

**Conclusion:** Overall, this new method allows to automatically and accurately segment the LC and offers the opportunity to measure its integrity both in terms of intensity and volume. This is of importance since these two metrics may offer complementary insights into LC integrity, as evidenced by the differing influences of demographic factors on LC intensity and volume.

## 1. Introduction

The locus coeruleus (LC) is a small (12–17 mm long and 2.5 mm wide) cylindrical nucleus located in the brainstem, serving as the principal site for the synthesis and release of norepinephrine (Fernandes et al., 2012; German et al., 1988; Poe et al., 2020). This pair of nuclei projects extensively throughout the brain and plays a crucial role in a myriad of functions including attention, stress responses, anxiety, neuroinflammation, arousal, sleep–wake cycle and memory (David and Malhotra, 2022; James et al., 2021; Morris et al., 2020; Poe et al., 2020; Van Egroo et al., 2022). Importantly, impairment of the LC has been linked to a range of neurological and psychiatric conditions, such as Alzheimer’s disease (AD), Parkinson’s disease (PD), and mood disorders (Chen et al., 2022; Gesi et al., 2000; Matchett et al., 2021; Poe et al., 2020). For instance, histopathological studies have described the presence of neurofibrillary tangles (NFTs) - a hallmark lesion of AD - in the LC at very early stages of the disease, preceding their accumulation in cortical regions (Attems et al., 2007; Braak et al., 2011; Duyckaerts et al., 2009; Grudzien et al., 2007). In addition, a study reported neuronal loss in the LC of around 30% in patients with mild cognitive impairment (MCI) and 55% in patients with AD dementia, and the number of neurons correlated significantly with cognitive performances (episodic memory, working memory, visuo-spatial ability) measured ante-mortem (Kelly et al., 2017).

All these observations stressed the need to measure LC integrity *in vivo* in humans and recent technical developments in MRI offered this opportunity. More precisely, neuromelanin (NM) – a paramagnetic pigment present in the catecholaminergic neurons of the LC – appears hyperintense on T1-weighted Turbo Spin Echo MRI sequences potentially due to its paramagnetic properties and high affinity for metal ions like iron and copper (Keren et al., 2015, 2009; Sasaki et al., 2006; Zucca et al., 2006). This approach, which allows visualization and estimation of LC integrity, has been increasingly used in recent years. Using this method, researchers have explored the role of the LC in the pathogenesis of neurodegenerative diseases, confirming histopathological findings and paving the way for the development of new sensitive imaging biomarkers (Chen et al., 2022; Galgani et al., 2022, 2020; Liu et al., 2017). Indeed, NM-MRI studies consistently reported lower LC signal intensity in patients with AD (Betts et al., 2019; Dordevic et al., 2017; Lagarde et al., 2024; Olivieri et al., 2019) or PD (García-Lorenzo et al., 2013; Ohtsuka et al., 2014; Schwarz et al., 2017; Wang et al., 2018) compared to healthy controls. In addition, LC intensity correlates with tau burden measured either in fluids or by PET in AD, reflecting the potential importance of these nuclei in AD pathophysiology (Betts et al., 2019; Dahl et al., 2022; Jacobs et al., 2022; Van Egroo et al., 2023). However, studies examining the impact of age on LC integrity yield mixed results, with some reporting significant effects (Betts et al., 2017; Clewett et al., 2016; Liu et al., 2019; Shibata et al., 2006) and others finding none (Al Haddad et al., 2023; Giorgi et al., 2022). The same conclusion applies to the effects of sex, with studies repotting LC signal differences between men and women (Bennett et al., 2024; Clewett et al., 2016) while others did not (Calarco et al., 2022; Jacobs et al., 2021; Shibata et al., 2006). To our knowledge, only one study investigated the effects of education on LC integrity and found no significant association (Clewett et al., 2016). Segmenting the LC is challenging due to its small size, even in ultra-high-resolution images. To circumvent the time-intensive nature of manual segmentation and the anatomical expertise it requires, semi or fully automated methods have been developed to assess LC integrity. Importantly, these techniques most often rely on a single template built in a standardized space and/or depend on a number of voxels to be accounted that is defined a priori (Betts et al., 2017; Cassidy et al., 2022; Clewett et al., 2016; Dahl et al., 2019; García-Lorenzo et al., 2013; Jacobs et al., 2022). Thus, these algorithms make it impossible to perform direct volumetric analyses and do not properly account for inter-individual anatomical variability.

The primary goal of this study was to develop a fully automated, multi-atlas segmentation method designed to replicate manual segmentation. This method enables direct segmentation of the LC in native space, accommodating inter-individual anatomical variability and facilitating volumetric analyses. To that end, we used the *Automatic Segmentation of Hippocampal Subfields* (ASHS) software (Yushkevich et al., 2015). Manual segmentations performed on ultra-high resolution NM images in 30 cognitively unimpaired (CU) older adults were fed into the ASHS training pipeline to generate a new atlas (ASHS-LC). After validation, this method was applied to an independent group of 71 CU to investigate the effects of age, sex and education on LC intensity and volume.

## 2. Materials and methods

### a. Participants

One hundred and four participants from the Age-Well randomized clinical trial of the Medit-Ageing European Project were included in the present study (Poisnel et al., 2018). Participants were recruited from the general population from November 2016 until March 2018, older than 65 years, native French speakers, retired for at least 1 year, educated for at least 7 years, and performed within the normal range on standardized cognitive tests. They did not show evidence of a major neurological or psychiatric disorder (including alcohol or drug abuse), history of cerebrovascular disease, presence of a chronic disease or acute unstable illness, and current or recent medication usage that may interfere with cognitive functioning. Data used in this study were acquired during the second post-intervention visit which took place from October 2020 until November 2022 (between 2.5 and 3 years after the end of the intervention). Three participants were not included in the analyses due to poor quality scans or discomfort during the MRI session (no images were acquired), resulting in 101 participants. Demographics of the cohort are reported in Table 1.

**Table 1:** Demographics of the population.

|  | Entire sample | Atlas set | Analysis set |
| --- | --- | --- | --- |
| N | 101 | 30 | 71 |
| Age (years ± SD) | 73.8 (± 6.6) | 73.9 (± 3.3) | 73.7 (± 3.7) |
| Gender (F,M) | 58/43 | 16/14 | 42/29 |
| Education (years ± SD) | 13.3 (± 3.0) | 12.9 (± 3.0) | 13.5 (± 3.0) |

The Age-Well randomized clinical trial was approved by the local ethics committee (Comité de Protection des Personnes Nord-Ouest III, Caen, France; trial registration number: EudraCT: 2016–002,441–36; IDRCB: 2016-A01767–44; ClinicalTrials. gov Identifier: NCT02977819) and all participants gave written informed consent prior to the examinations.

### b. MRI data acquisition

Each subject underwent an MR scan at the CYCERON center (Caen, France) using a 3T GE Healthcare SIGNA Premier scanner. First, T1-weighted BRAVO structural images (referred to as T1w in the rest of the manuscript) were acquired (Repetition Time (TR) = 7.2 ms; Echo Time (TE) = 2.9 ms; flip angle = 6◦; 180 slices; slice thickness = 1 mm; Field of View (FoV) = 256 × 256 mm2; matrix = 256 × 256; in-plane resolution = 1 × 1 mm2; acquisition time = 5 min28). In addition, 3 ultra-high resolution T1-weighted TSE images with additional magnetization transfer contrast (referred to as T1-MT in the rest of the manuscript) were acquired aligned with the anatomic axis of each participant’s brainstem (Repetition Time (TR) = 660 ms; Echo Time (TE) = 12 ms; slice thickness = 3 mm; Field of View (FoV) = 216 × 216 mm^2^; matrix = 512 × 512; in-plane resolution = 0.42 × 0.42 mm^2^; number of signal averages (NSA): 2; acquisition time = 4 min17). The three scans were then coregistered and averaged using SPM12 as well as resampled to a resolution of 0.3 × 0.3 × 0.75mm^3^ with a *Sinc* function using c3d.

### c. LC segmentation

#### i. Manual segmentation

To build the ASHS-LC atlas, 30 subjects were randomly selected (Atlas set – see Table 1) and the LC were manually segmented on each T1-MT image (in native space) using ITK-SNAP (http://www.itksnap.org/) (Yushkevich et al., 2006). Based on previous publications, the uppermost LC slice in the axial plane was identified immediately following the lower boundary of the interpeduncular fossa at the level of the inferior colliculus. The LC segmentation proceeded from the dorsal to ventral direction within the axial plane, usually extending to the lowest slices of the 4th ventricle, which is anatomically located at the level of the superior cerebellar peduncle (Betts et al., 2017; Priovoulos et al., 2018). Note that both left and right LC were segmented using a single label. All manual segmentations were made by the same rater (TB).

#### ii. Automatic segmentation using ASHS

The T1w MRI, T1-MT MRI together with the bilateral manual segmentations in the space of the T1-MT MRI were fed into the ASHS training pipeline to generate the ASHS-LC atlas (Figure 1 A). The ASHS pipeline is publicly available (https://www.nitrc.org/projects/ashs) and was described in details in Yushkevich et al., (2015) and Xie et al., (2019). Briefly, the training pipeline summarizes in the following steps:

- An unbiased whole brain population template is built using the T1w MRI of all the subjects.
- The region of interest (ROI) was identified by averaging the corresponding manual segmentations that are warped to the space of the template.
- Each T1-MT MRI and the corresponding segmentation were warped to the space of the template and cropped around the ROI.
- Pairwise registrations between all the subjects were performed between the warped and cropped scans.
- Label fusion was performed for each atlas in its native space using the rest of the atlases as candidates.
- AdaBoost classifiers were trained to learn the systematic error between the automatic segmentation and the manual segmentations.

**Figure 1:**
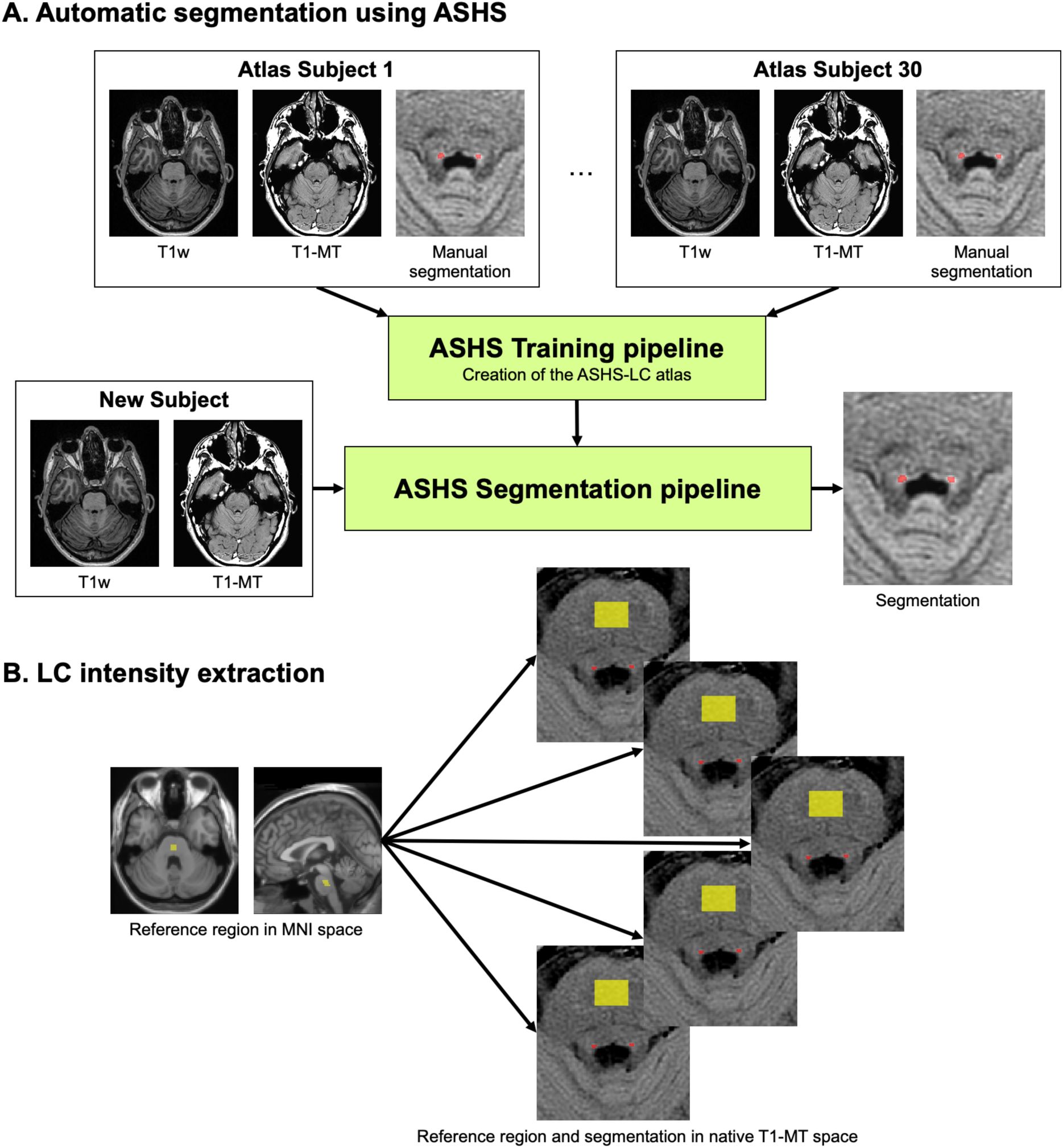
Overview of the processing pipeline. A. Graphical illustration of the training and segmentation pipelines in ASHS. B. Intensity extraction in the LC and reference ROIs.

Once trained, the segmentation pipeline with the ASHS-LC atlas was applied to the remaining 71 subjects (Analysis set – see Table 1). Briefly, the segmentation pipeline summarizes in the following steps:

- The ROI around the LC is identified in the target T1-MT image by registering to a whole-brain template generated in the training pipeline.
- For each target ROI, the corresponding ROIs in the atlas set are registered to it using Greedy (Venet et al., 2021).
- Atlas labels are then warped to the target ROI and combined using the joint label fusion algorithm (Wang and Yushkevich, 2013).
- The process is repeated in a bootstrapping fashion, where the initial segmentation of the target structures is used to initialize affine alignment between the atlas and target ROIs. This bootstrapping results in fewer failed atlas-to-target registrations and better over-all segmentation accuracy.

Resulting segmentations were all visually checked and 6 were manually edited. Segmentation time on a single core takes about 20 minutes for a subject.

### d. LC intensity / volume extractions

The ASHS segmentation produces 3 output files: *heur*, *corr_nogray* and *corr_usegray* (Xie et al., 2019). In this study, the *heur* segmentations were used.

LC intensity (LC_int_) was calculated as the average signal within the LC ROI (segmented manually or automatically) extracted directly from the T1-MT scan in native space. LC intensity was then normalized to a reference region located in the rostral pontomesencephalic area (Betts et al., 2017; García-Lorenzo et al., 2013). The reference region (729mm^3^) was originally segmented manually in MNI space and was then transformed to T1-MT scan native space using SPM12. The average signal was calculated within the reference region ROI (reference_int_) and was used to normalized the LC intensity using the following formula: LC_int_ratio_ = LC_int /_ reference_int_ (Figure 1 B).

To compensate for interindividual variability in head size, total intracranial volume (TIV, calculated using SPM12 by summing the volumes of the gray matter, white matter and cerebrospinal fluid) was regressed out from raw LC volumes (LC_vol_). From this point onward, normalized volumes will be referred to as LC_vol_TIV_ throughout the manuscript

### e. Statistical analyses

The accuracy of the ASHS segmentation was evaluated in subjects from the altas set (n=30) by performing five leave-six-out cross-validation experiments. In brief, five distinct atlases were generated by excluding each time six subjects from the total of 30. For a given subject, the ASHS segmentation that was performed using the atlas in which this subject had been excluded was compared to the corresponding manual segmentation. Dice similarity coefficients (DSC; Dice, 1945) were calculated to estimate the overlap between the automatic segmentations and the corresponding manual segmentations for each image. Then, for both intensity and volume, ASHS and manual segmentations were compared by: i) computing intraclass correlation coefficients [ICC(2,1) (Koo and Li, 2016)], ii) computing paired t-tests and iii) generating Bland-Altman plots to visualize the agreement between ASHS and manual segmentations. Lastly, correlations between automatic and manual segmentation were assessed for intensity and volume. All analyses were performed on raw, unnormalized measurements.

The effects of age, sex and education on LC integrity were investigated in subjects from the analysis set (n=71) using both intensity (LC_int_ratio_) and volume (LC_vol_TIV_) measurements generated with ASHS. The effects of age were assessed using partial correlations, with sex, education and intervention group as covariates. The effects of sex were evaluated using ANCOVAs, controlling for age, education and intervention group while the effects of education were analyzed using partial correlations, with age, sex and intervention group as covariates.

Lastly, the association between LC intensity and volume was assessed in the atlas set using manual and automatic segmentation (separately) as well as in the analyses set using automatic segmentation (since manual segmentation was not available in this group).

Results were considered significant when p < 0.05. All statistical analyses were performed using R version 4.1.2 with the stats, QuantPsyc, effsize and irr packages.

## 3. Results

### a. Accuracy of the ASHS segmentation

As illustrated in Figure 2, ASHS and manual segmentations showed strong overlap with a DSC of 0.83 ± 0.04.

**Figure 2:**
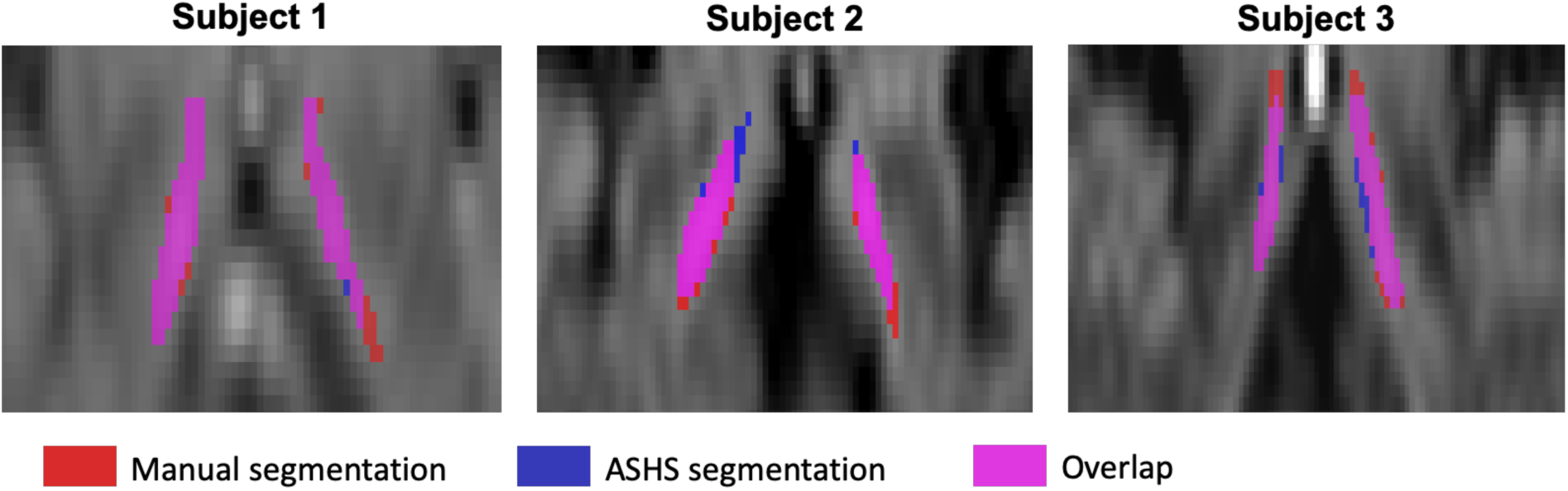
Example of manual and ASHS segmentations on 3 subjects from the atlas set.

ICCs demonstrated excellent reliability for intensity (ICC = 0.99), whereas reliability for volume was lower (ICC = 0.43). Paired t-tests revealed significant differences between intensity and volume measures from ASHS vs manual segmentations (p < 0.001 in each case). The effect was negligible for intensity (mean difference = -2.77, Cohen’s d = -0.02) but large for volume (mean difference = 3.79, Cohen’s d = 0.99). As illustrated on the Bland-Altman plots (Figure 3 A), ASHS slightly overestimates intensity and underestimates volume. Lastly, measures from manual and ASHS segmentation were significantly correlated for both intensity (β = 0.99, p < 0.001) and volume (β = 0.70, p < 0.001).

**Figure 3:**
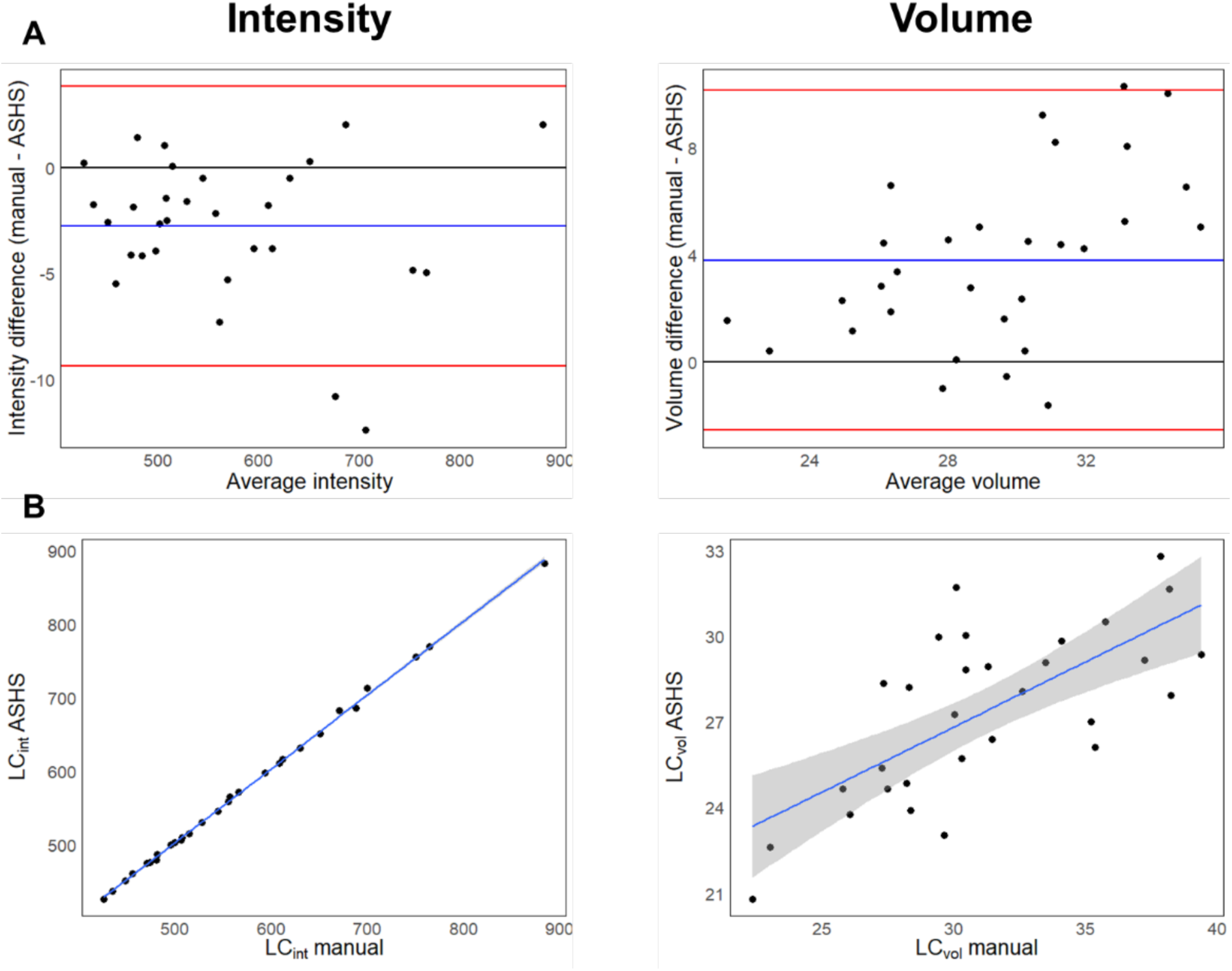
Comparison between manual and ASHS segmentation. A. Bland-Altman plots for intensity and volume. B. Correlations between manual and ASHS segmentation for intensity and volume.

**Figure 4:**
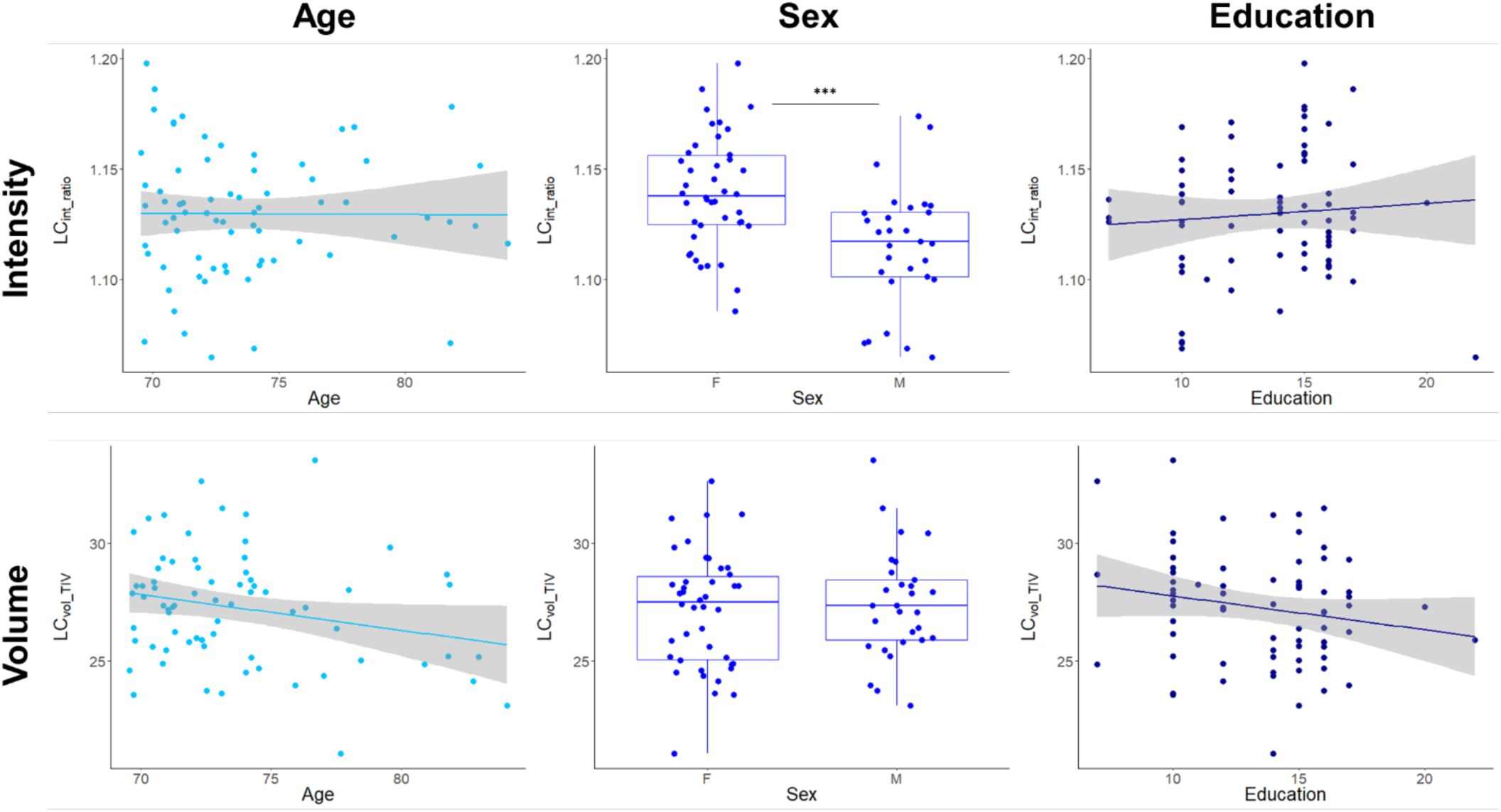
Effects of age, sex and education on LC integrity.

### b. Effects of age, sex and education on LC integrity

LC intensity was significantly higher in women than in men (Cohen’s d = 0.94, p < 0.001) while no associations with age (β = -0.0002, p = 0.98) or education (β = 0.11, p = 0.37) were found. In contrast, LC volume was not different between men and women (Cohen’s d = 0.10, p= 0.55) but was significantly negatively associated with age (β = -0.25, p = 0.04) and education (β = - 0.27, p = 0.04).

### c. Associations between LC intensity and volume

LC intensity and volume were not statistically correlated, neither using manual segmentation (atlas set: β = 0.05, p = 0.78) nor ASHS segmentation (atlas set: β = 0.11, p = 0.57; analysis set: β = -0.07, p = 0.55).

## 4. Discussion

In the present study, we first aimed to develop and validate a fully automated, multi-atlas segmentation method (ASHS-LC) for the locus coeruleus (LC) using ultra-high-resolution neuromelanin-sensitive MRI. This method aimed to address limitations of previous segmentation techniques, such as reliance on single templates and inadequate handling of inter-individual anatomical variability. Our findings demonstrated high accuracy of the ASHS-LC method, with strong agreement between manual and automated segmentations for intensity measures and moderate agreement for volume measures. A second aim was to investigate the effects of age, sex and education on LC intensity and volume. Our analysis showed differing impacts of demographic factors on LC intensity and volume.

### Accuracy of ASHS-LC Segmentation

The Dice similarity coefficient (DSC) of 0.83 indicates substantial overlap between manual and automated segmentations, exceeding previous reports using similar automated pipelines. Indeed, the DSCs for LC automatic segmentation reported in the literature were 0.40 (Ariz et al., 2019), 0.54-0.64 (Sibahi et al., 2023), 0.62-0.65 (Aganj et al., 2024) and 0.60-0.71 (Dünnwald et al., 2021). Intensity measures showed excellent reliability (ICC = 0.99), while volume measures exhibited lower reliability (ICC = 0.43). The systematic bias observed, with ASHS slightly overestimating intensity and underestimating volume, suggests that while intensity extraction is highly robust, volumetric measurements may still be influenced by segmentation inconsistencies. This discrepancy highlights the ongoing challenge of accurately capturing the small and anatomically variable LC structure, even with advanced segmentation algorithms. Nevertheless, ASHS-derived volumes demonstrated a strong correlation with manual volumes (β = 0.70, p < 0.001), suggesting that ASHS segmentation may serve as a reliable proxy.

### Effects of Age, Sex, and Education on LC Integrity

Our analysis revealed divergent effects of demographic factors on LC intensity and volume. LC intensity was significantly higher in women than in men. While several studies did not report sex differences on LC integrity (Calarco et al., 2022; Jacobs et al., 2021; Shibata et al., 2006), others did (Bennett et al., 2024; Clewett et al., 2016). However, these studies reported higher integrity in males than in females, either using NM-MRI (Clewett et al., 2016) or diffusion MRI (Bennett et al., 2024). Notably, these analyses included both young and older adult groups. Interestingly, Clewett et al. (2016) reported no significant sex differences in LC signal intensity when examining each age group separately. Regarding LC intensity, our analyses did not show significant associations with either age or education. In contrast, LC volume demonstrated significant negative associations with both age and education, but no differences between sexes. The age-related decline in LC volume is consistent with previous histopathological and imaging studies indicating neuronal loss in the LC with aging (Betts et al., 2017; Clewett et al., 2016; Liu et al., 2019; Shibata et al., 2006). Interestingly, a higher level of education was linked to smaller LC volume. This association might stem from increased stress and sleep disturbances that may be experienced by highly educated individuals, which can negatively impact LC integrity.

### LC Intensity and Volume Relationship

LC intensity and volume were not significantly correlated, regardless of whether manual or automated segmentation was used. This finding suggests that intensity and volume might reflect distinct neurobiological properties of the LC, with intensity potentially representing neuromelanin concentration and volume reflecting global structural integrity. These divergent measures highlight the importance of considering both metrics in future studies investigating LC integrity.

### Methodological Considerations and Future Directions

While ASHS-LC offers significant advantages, including reduced processing time and enhanced reproducibility, certain limitations must be acknowledged. The lower reliability of volume measures emphasizes the need for ongoing refinement of segmentation algorithms, particularly in regions with complex anatomical variability. Furthermore, the study’s cross-sectional design limits causal interpretations of observed associations. Future longitudinal studies are warranted to investigate the dynamic changes in LC integrity over time and their relationship with cognitive decline and neurodegenerative processes.

In conclusion, this study presents a validated automated pipeline for LC segmentation, enabling reliable extraction of LC intensity and volume from neuromelanin-sensitive MRI. The ASHS-LC atlas is planned for public release in the near future. The observed associations with demographic factors and the dissociation between intensity and volume measures underscore the complexity of LC structural and functional integrity. These findings pave the way for future research exploring the LC’s role in aging, neurodegeneration, and cognitive resilience.

## Data Availability

All data produced in the present study are available upon reasonable request to the authors

